# Association between Interictal Spike Rate and Seizure Frequency in a Large Epilepsy Cohort

**DOI:** 10.64898/2026.02.24.26346988

**Authors:** Erin C. Conrad, Ellie Chang, Kevin Xie, Carlos A. Aguila, Juri Kim, Haoer Shi, William KS Ojemann, Jin Jing, M. Brandon Westover, Saurabh R. Sinha, Brian Litt, Kathryn A. Davis, Quy Cao, Colin A. Ellis

## Abstract

**Background and Objectives:** Tracking and predicting seizure frequency in patients with epilepsy is important for prognostication and therapy management. Interictal spikes have been proposed as a biomarker of seizure burden, but their association with seizure frequency has not been well quantified across epilepsy subtypes. Our objective was to measure the association between spike rate and seizure frequency and how this varies by epilepsy subtype.

**Methods:** We studied 3,614 consecutive routine outpatient EEGs from 3,245 patients with epilepsy followed for a median of 2.8 years. A validated automated detector (SpikeNet2) estimated spike frequency. Validated large language models performed natural language processing on outpatient clinic notes to extract seizure frequency and epilepsy subtype. We measured Spearman correlation between spike frequency (spikes/hour) and seizure frequency (seizures/month) for all patients with epilepsy and for patients with generalized epilepsy, temporal lobe epilepsy, and frontal lobe epilepsy.

**Results:** Overall, spike frequency was modestly associated with seizure frequency (N = 3,245, ρ = 0.11 [95% CI 0.07-0.14], p < 0.001). Significant positive associations were observed in generalized epilepsy (N = 625, ρ = 0.23 [0.15-0.30], Bonferroni-adjusted p < 0.001) and temporal lobe epilepsy (N = 834, ρ = 0.12 [0.05-0.19], p = 0.0013), but not in frontal lobe epilepsy (N = 263, ρ = 0.11 [-0.02-0.24], p = 0.22). A mixed-effects model revealed a stronger spike-seizure association for clinic visits close in time to EEGs (OR=0.983 [0.967-0.999], p = 0.03), suggesting that spike rate may be a time-varying biomarker of seizure frequency.

**Discussion:** In this large outpatient cohort, higher interictal spike rates on routine EEG were associated with higher seizure frequencies, with the strongest relationship observed in generalized epilepsy. These associations support interictal spike rate as a quantitative EEG marker of seizure burden. Spike rate may have clinical utility for risk stratification at diagnosis and for monitoring longitudinal changes in seizure burden in response to therapy.

## INTRODUCTION

Epilepsy is characterized by recurrent unprovoked seizures. Accurate estimation of seizure frequency informs counseling, monitoring intensity, and therapy modifications. Clinically, seizure frequency is assessed through patient self-report at outpatient visits occurring every 3–12 months. This approach is limited by inaccuracy (patients are unaware of approximately half of their seizures) ^1^, and the delay imposed by the paroxysmal, infrequent nature of seizures, which requires months of observation before burden can be reliably estimated. This prolonged and imprecise process exposes patients to preventable morbidity. We need objective biomarkers of seizure frequency that can be measured at any time.

Interictal spikes are brief paroxysmal EEG waveforms associated with epilepsy diagnosis. Spikes occur much more frequently than seizures ^2^, making spike rate a promising continuous biomarker of epilepsy severity. Whether spike burden reflects seizure frequency has been examined in several prior studies, with mixed results ^3–8^. Prior inconsistent findings likely reflect several limitations of the existing literature. First, most studies enrolled tens to low hundreds of patients, limiting power to detect modest associations. Second, epilepsy’s syndromic heterogeneity may produce subtype-specific spike-seizure relationships that dilute aggregate findings. Third, prior work has largely treated spike burden as a static patient-level measure rather than examining whether it tracks within-individual changes in seizure frequency over time. Finally, scalable methods to quantify both spikes and seizure outcomes across large, heterogeneous cohorts have historically been lacking, limiting the generalizability of findings.

Advances in natural language processing (NLP) ^9^ and automated spike detection ^10^ now allow estimation of seizure and spike frequency at scale (**Fig. 1**). EEG-based biomarkers of seizure frequency are attractive because scalp EEG is inexpensive and widely available, and emerging subgaleal EEG devices will soon enable chronic monitoring ^11^. We applied validated large language models (LLM) ^9,12^ and an automated spike detector ^10^ to electronic medical records and routine EEGs from a large epilepsy cohort to quantify the association between spike rate and seizure frequency, examine whether this association varies across epilepsy subtypes, and test whether it reflects within-individual changes in seizure burden over time.

**Figure 1.**
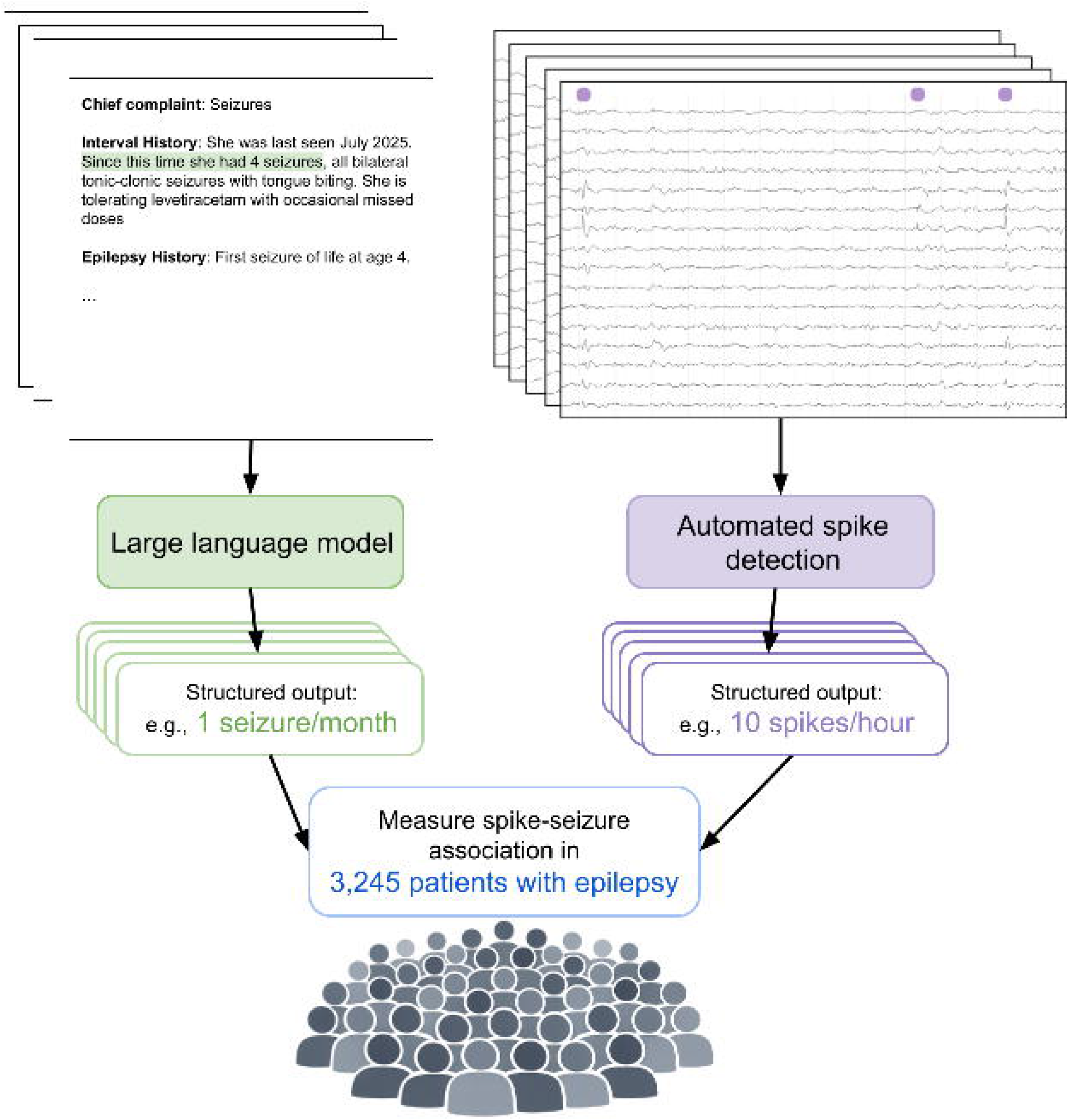
Scalable extraction of seizure frequency and spike rate from routine clinical data. Seizure frequency and interictal spike rate were quantified at scale using two parallel automated pipelines applied to electronic medical records from 3,245 patients with epilepsy. (Left) A large language model extracted seizure frequency from free-text neurology clinic notes, producing structured output (e.g., seizures/month) for each outpatient visit. Highlighted text illustrates a fictional interval history (shown for example) from which seizure frequency was extracted. (Right) An automated spike detector was applied to routine outpatient EEGs, identifying interictal spikes (purple markers) and producing a structured spike rate per recording (e.g., spikes/hour). Outputs from both pipelines were linked at the patient level to quantify the association between spike rate and seizure frequency across the cohort.

## METHODS

### Standard protocol approvals, registrations, and patient consents

We conducted a retrospective cohort study of patients evaluated at the Penn Epilepsy Center at the University of Pennsylvania between 2014 and 2024. The study was approved by the University of Pennsylvania Institutional Review Board (protocol # 835008) with waiver of informed consent.

### Participants

Sample size was determined by the available clinical database. We identified patients who had at least one routine outpatient EEG (duration <4 hours) and at least one outpatient clinic visit with a recorded seizure frequency. Patients were included if they carried an LLM-confirmed epilepsy diagnosis (see below). Patients without an LLM-confirmed epilepsy diagnosis, documented seizure frequency, or EEG were excluded. We included patients regardless of age or sex, though typically only adult patients are seen at our center. As this was a retrospective study with no follow-up, loss to follow-up was not applicable.

### Extraction of Seizure Frequency and Epilepsy Phenotype

Clinical data were extracted from outpatient epilepsy clinic notes using validated natural language processing (NLP) tools previously described ^9,12–14^. Seizure frequency (seizures/month) was extracted using fine-tuned Bio_ClinicalBERT and RoBERTa transformer models trained on 700 annotated clinical notes, with performance comparable to human interrater agreement (model F1 = 0.85, human F1 = 0.85). Patients documented as seizure-free at a visit were assigned a frequency of zero for that visit; visits without a documented frequency were excluded from patient-level estimates. The proportion of clinic visits with documented seizure frequency varied across patients (**Table 1**). Patient-level mean seizure frequency was computed by averaging across all visits.

**Table 1.** Cohort characteristics. Seizure frequency reflects the mean number of reported seizures per month across clinical visits; seizure frequency was considered documented at a visit if a numeric frequency was recorded or if the patient was explicitly noted to be seizure-free since the last visit. Spike rate represents the mean automated spike-detection rate across outpatient routine EEGs. Follow-up duration is the time between a patient’s first and last outpatient clinic visit. Epilepsy diagnosis, epilepsy subtype, and mean monthly seizure frequency were extracted from clinic notes using large language models. Age, sex, number of clinic visits, follow-up duration, and the presence or absence of reported spikes were derived from structured variables in the electronic medical record. Values are median (IQR) unless otherwise noted.

|  |  |
| --- | --- |
| Patients | 3245 |
| Age at first visit<br>Median (IQR) | 35.6 (25.1-52.6) |
| Sex<br>N (%) |  |
| Women | 1750 (53.9%) |
| Men | 1494 (46.0%) |
| Unknown/Other | 1 (0.0%) |
| Epilepsy subtype<br>N (% Epilepsy) |  |
| Temporal lobe | 834 (25.7%) |
| Frontal lobe | 263 (8.1%) |
| Generalized | 625 (19.3%) |
| Other | 1175 (36.2%) |
| Unknown | 348 (10.7%) |
| Number of clinic visits<br>Median (IQR) | 5.0 (2.0-10.0) |
| Years of follow-up<br>Median (IQR) | 2.8 (0.6-6.1) |
| % Visits with<br>documented seizure<br>frequency<br>Median (IQR) | 68.8% (50.0-100.0) |
| Number of EEGs<br>Median (IQR) | 1.0 (1.0-1.0) |
| Mean monthly<br>seizures across visits<br>Median (IQR) | 0.73 (0.00-5.07);<br>median CI [0.62-0.92] |
| Mean spikes/hour<br>across EEGs<br>Median (IQR) | 1.79 (0.64-4.79);<br>median CI [1.69-1.93] |
| EEGs with reported<br>spikes<br>N (% EEGs) |  |
| Present | 375 (10.4%) |
| Absent | 2683 (74.2%) |
| Unknown | 556 (15.4%) |
| Patients with reported<br>spikes<br>N (% patients) |  |
| Present | 355 (10.9%) |
| Absent | 2458 (75.7%) |
| Unknown | 432 (13.3%) |

Epilepsy diagnosis, focal versus generalized classification, and focal localization (temporal vs. frontal, the most common localizations in our cohort) were extracted using DeepSeek-R1, a large language model applied with few-shot prompting to outpatient clinic notes ^15^. In the original validation study ^12^, model performance on 309 expert-annotated notes was comparable to human interrater agreement for six-way epilepsy classification (focal, generalized, combined generalized and focal, unclassified/unspecified, uncertain if epilepsy, or non-epileptic seizure disorder; model F1 = 0.78, human F1 = 0.82) and all-way subtype classification (see **Supplemental Materials** for these classifications; model F1 = 0.63, human F1 = 0.72).

For primary analyses, patients were classified as having epilepsy if their six-way classification was focal, generalized, combined generalized and focal, or unclassified/unspecified. For subtype analyses, temporal lobe epilepsy and frontal lobe epilepsy were assigned according to their all-way localization; generalized epilepsy was assigned based on the six-way classification (excluding combined generalized and focal).

### EEG Acquisition and Spike Detection

Scalp EEGs were performed as part of routine clinical care using the standard 10-20 electrode system, sampled at 256 Hz. We included outpatient EEGs with duration less than 4 hours, excluding inpatient and ambulatory recordings. Recordings were converted to EDF format and preprocessed for spike detection by applying a 60 Hz notch filter, a 0.5 Hz high-pass filter, and resampling to 128 Hz.

Automated spike detection was performed using SpikeNet2 ^10^, a deep learning model based on a residual network architecture that outputs time-varying spike probabilities at 16 samples/second. In the original publication, the area under the receiver operating curve and area under the precision recall curve were 0.942 and 0.948, respectively, for event-level spike classification in the external validation set relative to expert consensus ^10^. Detections were defined as probabilities exceeding 0.46 (from the original publication), with detections within a one-second window clustered into a single event. Spike rate was defined as the number of detections per hour of EEG. Patient-level mean spike rate was computed by averaging across all qualifying outpatient EEGs.

Clinically reported spike presence or absence was extracted by parsing EEG reports for standardized phrases indicating epileptiform discharge status. For 556 of 3,614 EEGs (15.4%), reported spike status could not be determined due to report unavailability or deviation from the standard reporting template.

### Statistical Analysis

All analyses were conducted in MATLAB R2024a (MathWorks).

#### Descriptive and correlation analyses

We examined the association between patient-level mean spike rate and mean seizure frequency using Spearman correlation, overall and stratified by epilepsy subtype (temporal lobe, frontal lobe, generalized, selected as these were the most common in our cohort). Subtype-specific p-values were Bonferroni-corrected for three comparisons. Group differences in spike rate by reported spike presence/absence and by epilepsy subtype were assessed using Mann–Whitney U-tests and Kruskal–Wallis tests, respectively, with effect size estimated using Cliff’s δ and η^2^. As a sensitivity analysis to assess whether results were driven by zero-inflated data, we repeated the Spearman correlation analyses restricting to patients with non-zero spike rates and non-zero seizure frequencies.

#### Mixed effects model

To test the association between spike rate and seizure occurrence while accounting for the time-varying nature of EEG-visit observations, we cross-joined each patient’s outpatient EEGs with their outpatient clinic visits to create a table of all possible EEG-visit pairs. We restricted analysis to patients with a documented epilepsy subtype—temporal, frontal, or generalized—so that we could include the epilepsy subtype (12,281 pairs from 1,722 patients). For each pair we computed the signed temporal lag (visit date minus EEG date), its absolute value, and a direction indicator (visit after vs. before EEG). Spike rate was log-transformed given its right-skewed distributions and wide dynamic range (natural log plus an offset of 0.001 spikes/hour to accommodate zero values). We then fit logistic mixed effects models predicting whether the patient reported having seizures at the clinic visit (yes/no) — chosen over visit-specific seizure frequency because seizure occurrence was documented more consistently across clinic notes. Fixed effects included log spike rate, absolute lag, lag direction (visit after vs. before EEG), epilepsy subtype (temporal as reference), and interactions between log spike rate and absolute lag (testing whether spike rate is more predictive when EEG and visit are close in time) and between log spike rate and lag direction (testing whether predictive value differs for EEGs obtained before vs. after the visit). A patient-level random intercept accounted for within-patient correlation. Model fit with and without interaction terms was compared using a likelihood ratio test, testing the hypothesis that adding temporal interaction terms will increase model fit, indicating a stronger spike-seizure association for clinic visits close in time to EEGs. Model formulas are shown in **Table S1**. Confidence intervals were estimated via patient-level cluster bootstrap (5,000 iterations).

As a secondary analysis, we examined whether the spike–seizure association was stronger when clinic visits occurred closer in time to EEG acquisition, which would be expected if spike rates track fluctuating seizure burden within individuals over time. We stratified visits into tertiles by the minimum absolute temporal distance between each visit and any EEG recorded for that patient, and compared Spearman correlations between spike rate and seizure frequency for visits in the lowest versus highest tertile. The difference in correlations between the close-in-time and far-in-time visits was tested using a patient-level bootstrap procedure (**Supplemental Materials**).

### Data availability

Code is publicly available at https://github.com/erinconrad/seizure_severity. The codebase includes clinical variables and spike counts needed to replicate the analyses. Raw EEG data is available upon reasonable request with a data use agreement.

## RESULTS

### Cohort summary

Of 6,544 patients with EEG data in the Penn Epilepsy Center database, 1,917 were excluded because their EEG was not an outpatient routine recording of less than 4 hours, 679 were excluded without an LLM-confirmed epilepsy diagnosis, and 703 were excluded without a documented seizure frequency, yielding a final cohort of 3,245 patients with 3,614 EEGs (**Fig. S1**). Most patients had only a single EEG throughout their follow-up. Median follow-up from first to last clinic visit was 2.8 years (IQR 0.6–6.1). Across patients, a median of 68.8% (IQR 50.0–100.0%) of clinic visits had a documented seizure frequency. 355 patients (10.9%) had spikes reported on at least one EEG. Median [95% CI] monthly seizure frequency was 0.73 [0.62-0.92], and median spikes/hour was 1.79 [1.69-1.93] (**Table 1**).

### Spike rates by patient groups

Detected spike rates were higher in EEGs with clinically-reported spikes (median 10.39 [95% CI 7.83-11.39] spikes/hour) than without (1.46 [1.37-1.60] spikes/hour) (p < 0.001, Cliff’s δ=0.65; **Fig. 2A**). Spike rates differed across epilepsy subtypes, with the highest rates in generalized epilepsy (Kruskal-Wallis p < 0.001, η^2^≈0.014; **Fig. 2B**).

**Figure 2.**
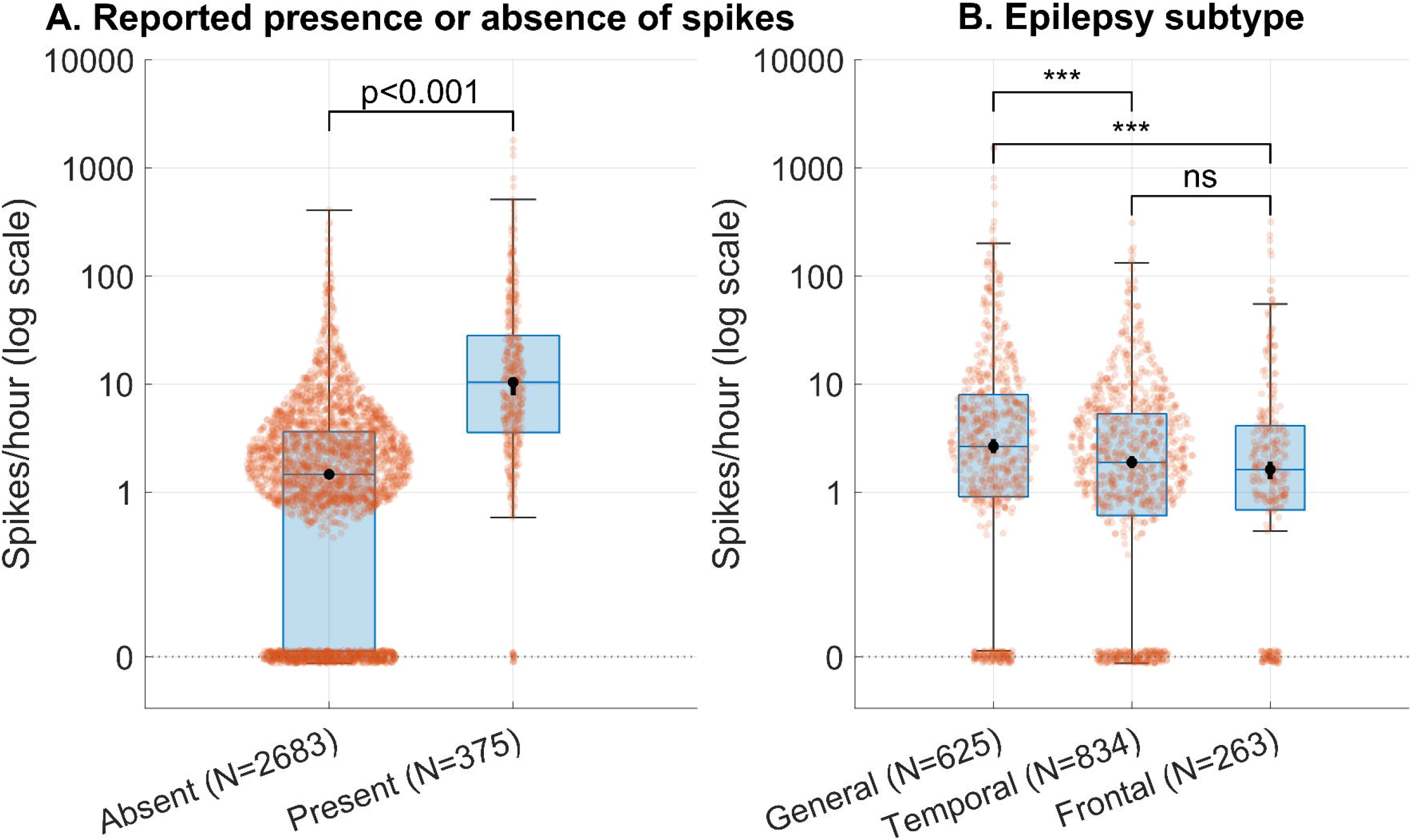
Spike rates across EEGs and cohorts. **A:** EEG-level spike rates detected by SpikeNet2 (spikes/hour, log□□ scale) for outpatient routine EEGs stratified by whether the clinical EEG report indicated “present’’ or “absent’’ epileptiform discharges. Red points represent individual EEGs, overlaid on boxplots (boxes indicate the interquartile range (IQR); center lines indicate the median; whiskers extend to 1.5×IQR). Black circles and vertical lines overlying the median represent 95% confidence intervals for the median derived from bootstrapping (5,000 iterations). The horizontal dotted line at 0 denotes EEG with a detected spike rate of 0, arbitrarily offset on the log scale from EEGs with non-zero spike rates. Most EEGs were less than one hour, and so spike rates between 0 and 1 per hour were not possible for most EEGs. For 556/3,614 EEGs, we could not automatically determine the presence or absence of spikes from the report. **B:** Per-patient mean SpikeNet2 spike rates (spikes/hour, log□□ scale) stratified by epilepsy subtype (generalized, temporal lobe, frontal lobe). A Kruskal–Wallis test showed overall group differences, with Bonferroni-corrected pairwise Mann–Whitney U-tests shown above comparisons. *** = Bonferroni-corrected p < 0.001. ns = Bonferroni-corrected p > 0.05. Generalized epilepsy demonstrated higher spikes/hour (median 2.65 95% CI [2.26-3.15]) than temporal (1.89 [1.66-2.16]; Bonferroni-adjusted p < 0.001) and frontal (1.62 [1.30-1.97]; p < 0.001). The temporal versus frontal comparison was not significant (p = 1.00).

### Spike rate and seizure frequency

Spike rate and seizure frequency were positively correlated across all epilepsy patients (N=3,245, ρ=0.11 [95% CI 0.07-0.14], p < 0.001). Subtype-specific correlations were significant for generalized epilepsy (N=625, ρ=0.23 [0.15-0.30], Bonferroni-adjusted p < 0.001) and temporal lobe epilepsy (N=834, ρ=0.12 [0.05-0.19], p = 0.0013), but not frontal lobe epilepsy (N=263, ρ=0.11 [-0.02-0.24], p = 0.22; **Fig. 3**). When restricting to patients with non-zero spike rates and seizure frequencies, results were similar, although the temporal epilepsy correlation was no longer significant in this subgroup (Bonferroni-adjusted p = 0.06), although the magnitude was similar (ρ=0.10 [0.02-0.19]; **Fig. S2**). Patients with spikes on at least one EEG had higher mean seizure frequencies (**Fig. S3**).

**Figure 3.**
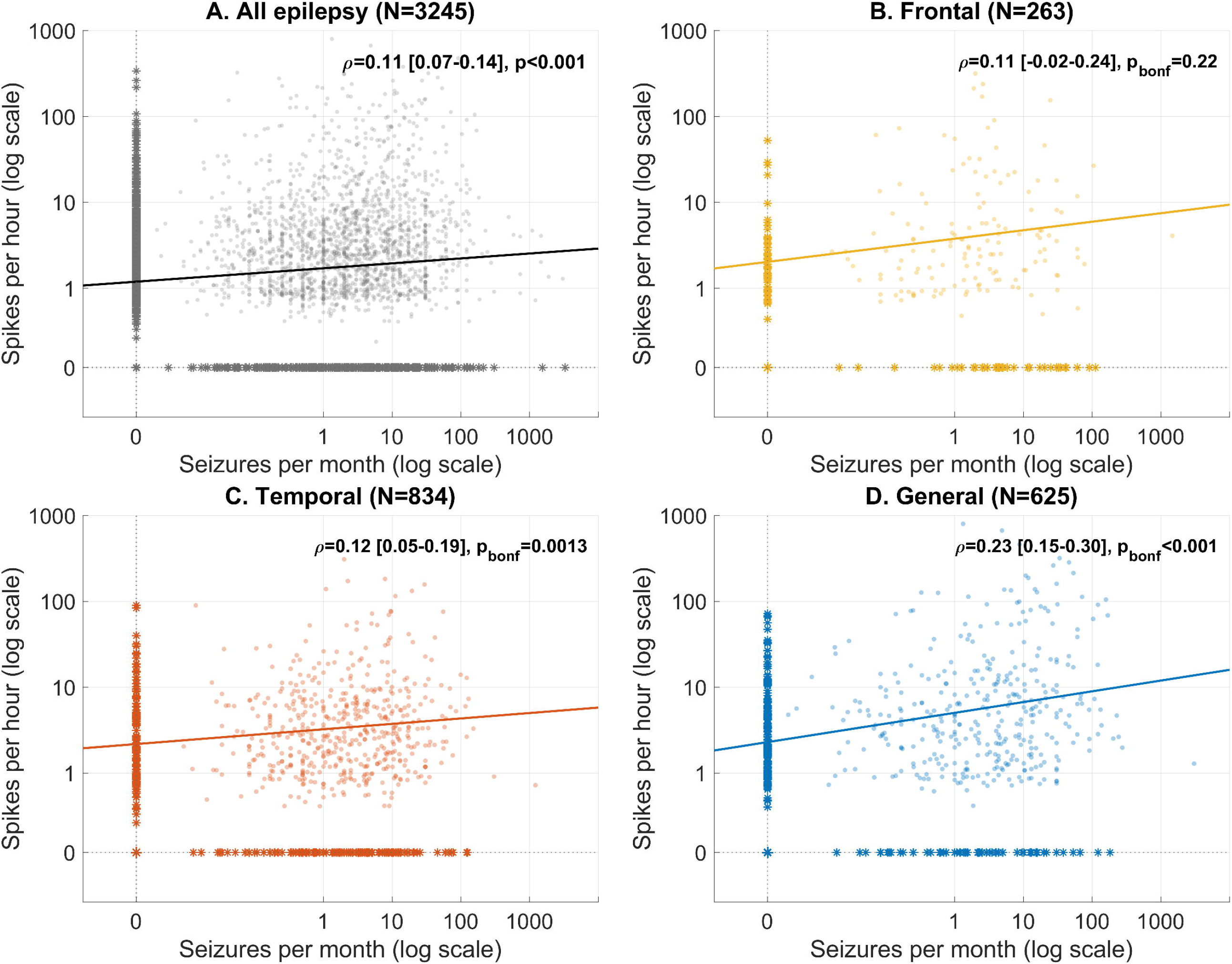
Relationship between spike rate and seizure frequency. **A-D**: Scatterplots showing the relationship between mean seizure frequency (seizures/month, log□□ scale) and mean spike rate (spikes/hour, log□□ scale). Each panel shows individual patients, fitted regression lines (for non-zero points), and Spearman correlation coefficients with associated 95% confidence intervals (obtained via Bootstrapping) and p-values (Bonferroni-corrected for subtype panels). The horizontal line at 0 spikes/hour with asterisks indicates patients with a mean spike rate of 0, and the vertical line at 0 seizures/month with asterisks indicates patients with a mean seizure frequency of 0 (arbitrarily offset from the rest of the data on the log scale). **A** shows results for all patients with epilepsy, **B** shows results for frontal lobe epilepsy, **C** shows results for temporal lobe epilepsy, and **D** shows results for generalized epilepsy.

### Mixed effects model to estimate time-varying spike-seizure association

Seizure frequency varies over time within individuals, and we hypothesized that spike rates track this variability, predicting a stronger spike-seizure association for clinic visits close in time to EEGs. To test this, we fit logistic mixed effects models on all EEG-visit pairs for patients with known epilepsy subtype (N=12,281 pairs, 1,722 patients), with interaction terms allowing the spike-seizure association to vary with the temporal distance between EEG and visit (**Fig. S4**). A likelihood ratio test confirmed that these interactions jointly improved model fit over a model without them (χ^2^(2), p < 0.001).

Higher spike rates were associated with higher odds of reporting seizures at a clinic visit (OR=1.10 [95% CI 1.05-1.16], p < 0.001; **Fig. 4A**), implying that a 2.7-fold increase in spike rate is associated with 10% higher odds of seizure reporting at a clinic visit. The spike-seizure association attenuated with greater EEG-visit distance, although the effect was small (OR=0.983 [0.967-0.999], p = 0.03): e.g., the model-predicted OR for spike rate was 1.10 at a 6-month lag, 1.07 at 2 years, and 1.03 at 4 years (**Fig. 4B**). Spike rates from EEGs obtained before versus after a clinic visit were similarly associated with seizure occurrence (interaction OR=1.020 [0.965-1.072], p = 0.49). Visits occurring after the EEG had a lower baseline odds of seizure reporting (OR=0.39 [0.32-0.48], p < 0.001), consistent with gradual clinical improvement over time (**Fig. S4**). Compared with temporal lobe epilepsy, generalized epilepsy had a lower baseline odds of seizure reporting (OR=0.49 [0.35-0.69], p < 0.001), while frontal lobe epilepsy did not differ significantly (OR=0.69 [0.43-1.09], p = 0.11). Our secondary analysis also found that spike-seizure correlations were stronger for clinic visits close in time to the EEG (**Fig. S5**). Together, these results confirm a positive spike rate–seizure association and suggest it is strongest when the EEG is obtained close to the clinic visit, consistent with spike rates tracking within-individual seizure burden over time.

**Figure 4.**
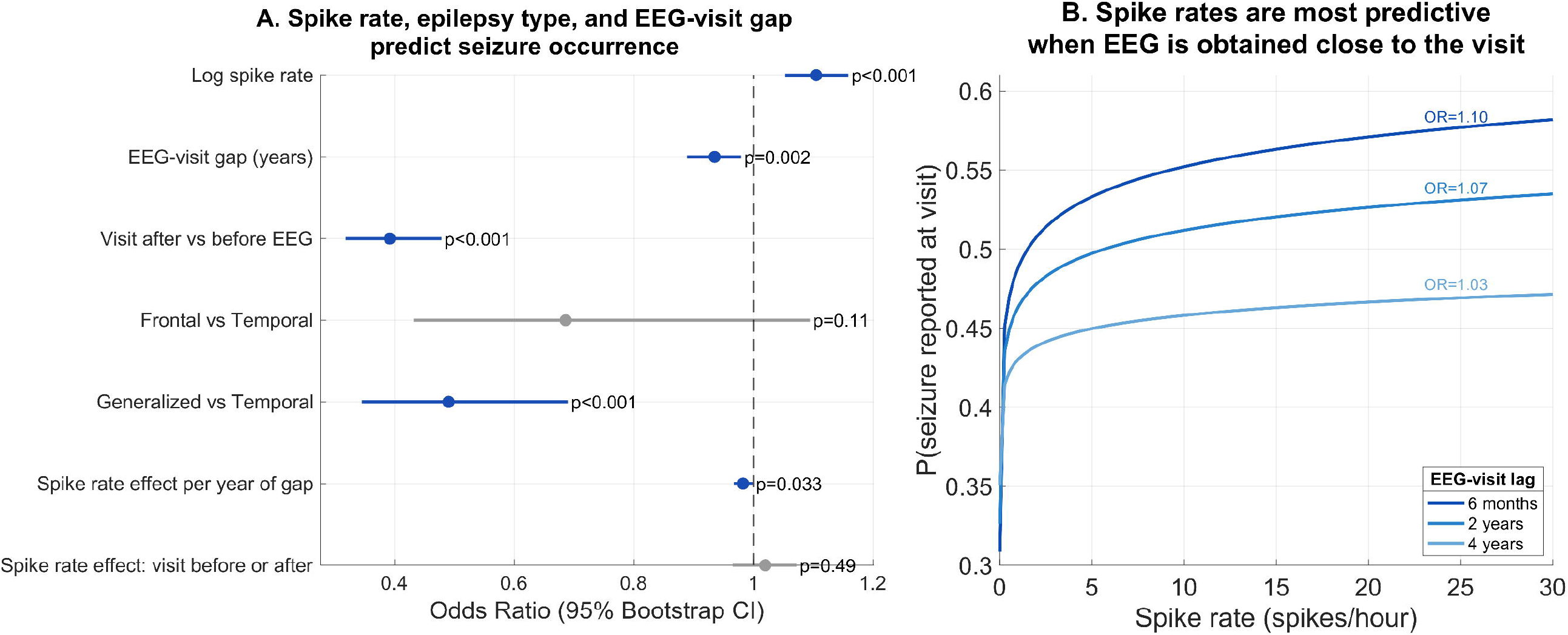
Spike rate predicts seizure occurrence, with attenuation over longer EEG-visit intervals. **A:** Forest plot of odds ratios (ORs) from a logistic mixed-effects model predicting whether a patient reported seizures at a clinic visit, fit on all EEG–visit pairs for patients with a known epilepsy subtype (N=12,281 pairs, 1,722 patients; patient-level random intercept). ORs with 95% bootstrap confidence intervals are shown; terms reaching significance (p<0.05) are shown in blue, non-significant terms in gray. Epilepsy subtype effects are referenced to temporal lobe epilepsy. **B:** Model-predicted probability of seizure occurrence as a function of spike rate, shown separately for three example EEG–visit lags (6 months, 2 years, 4 years), assuming a visit occurring after the EEG and temporal lobe epilepsy subtype. Annotated ORs reflect the per-unit-log spike rate effect at each lag distance. The attenuation of the spike rate effect with increasing lag is captured by the interaction term (Log spike rate × Absolute lag); see panel A.

## DISCUSSION

In this large outpatient cohort, higher interictal spike rates were associated with higher seizure frequencies, with the strongest associations in generalized and temporal lobe epilepsy. A logistic mixed effects model confirmed this association while accounting for repeated within-patient observations, and found that predictive value was greatest when EEGs were obtained close in time to clinic visits — consistent with spike rates tracking time-varying seizure burden. These findings were enabled by combining large language model–based chart extraction with automated EEG spike detection, which permitted analysis of a cohort an order of magnitude larger than prior work in this area.

Prior studies have reported mixed results ^3–8^, with inconsistency likely reflecting limited sample sizes, heterogeneous cohort composition, and reliance on static patient-level measurements that cannot capture within-individual variability. Our mixed effects model tested whether spikes behave as a time-varying biomarker rather than a fixed trait. The significant lag interaction supports this interpretation: predictive value attenuated as EEG-visit distance increased, consistent with spike rates tracking a dynamic rather than static disease state, although the absolute effect sizes were modest (OR=1.10 at 6 months, declining to 1.03 at 4 years).

Chronic intracranial EEG studies have demonstrated that reductions in spike rates track improvements in seizure control, but these are restricted to patients with drug-resistant focal epilepsy ^16,17^. Our findings support the plausibility of spike rate on scalp EEG as a dynamic biomarker, and motivate longitudinal studies pairing repeated or longitudinal scalp or subgaleal EEGs with prospective seizure outcome data.

The stronger spike-seizure association observed in generalized epilepsy may reflect greater biological relevance or improved detectability of generalized compared to focal spikes ^18^. The significant association in temporal lobe epilepsy is consistent with prior work and reinforces that spike burden reflects disease severity in focal as well as generalized syndromes. The absence of a significant association in frontal lobe epilepsy may reflect the limitations of scalp EEG for detecting frontal lobe activity ^19^, and the smaller sample size in that subgroup.

A central methodological contribution of this study is the demonstration that LLM-based extraction of seizure frequency from clinical notes and automated EEG spike detection can be combined to enable large-scale quantitative analysis of a question that has historically required labor-intensive manual review. Prior studies have typically enrolled tens to low hundreds of patients ^3–8^; our cohort of 3,245 represents a scale that dramatically improves the power to detect modest effect sizes in the setting of noisy clinical data, and the precision available for subgroup analyses. This pipeline is generalizable to other EEG biomarkers and clinical outcomes without prospective data collection.

### Limitations

Automated seizure frequency extraction and spike detection introduce measurement noise, as reflected by non-zero spike rates in some EEGs without clinically reported spikes. Seizure frequency was extracted from clinic notes, which reflect patient recall and may undercount events. Most patients contributed a single EEG, precluding a true longitudinal design, and the model may incompletely account for longitudinal changes in seizure burden — driven, for example, by regression to the mean or treatment response — that could spuriously drive the observed time-varying association. We lacked systematic medication data, precluding analysis of whether antiseizure medication changes mediated the attenuation of the spike-seizure association with longer lag times. We also could not account for modulators of spike rate including sleep deprivation, alcohol, or concurrent illness, which may have introduced noise into spike rate estimates. Unmeasured time-varying confounders would be expected to attenuate the observed association toward the null — for example, a patient who missed medications may have had more seizures before their clinic visit but been adherent at the time of EEG, dissociating the seizure and spike measurements and making our effect estimates conservative.

The majority of patients had no detectable spikes on routine outpatient EEG, consistent with prior literature reporting the sensitivity of spikes on EEG ^20^, implying that the observed association reflects both the difference in seizure burden between patients with and without spikes and variation in spike rate and seizure burden among those with detectable spikes. Approximately one-third of visits had no reported seizure frequency and were excluded from analysis; if missingness was systematically related to seizure burden, this could bias our results. Routine outpatient EEGs are brief snapshots that may miss spikes in patients with low spike rates or state-dependent spiking, potentially misclassifying some patients as spike-negative.

### Conclusions

Higher interictal spike rates on routine outpatient EEG were associated with higher seizure frequencies across a large and heterogeneous epilepsy cohort, with a stronger association when EEGs are obtained proximate to clinical assessment. The scalable pipeline combining LLM-based chart extraction with automated spike detection demonstrates a generalizable approach to EEG biomarker research. These results support spike burden as a promising noninvasive biomarker of seizure frequency and motivate chronic EEG studies to determine whether serial spike rates can guide individualized epilepsy management.

## Data Availability

Participant-level data of seizure frequencies and spike rates needed to replicate the analyses are publicly available in the link provided. Raw EEG data will be made available to researchers upon reasonable request after completing a data use agreement between institutions.

https://github.com/erinconrad/seizure_severity

