## Supplementary Materials for "Association between Interictal Spike Rate and Seizure Frequency in a Large Epilepsy Cohort"

**SUPPLEMENTAL METHODS**

*Details of extraction of seizure frequencies and epilepsy types using natural language processing*

The NLP development and validation of the NLP tools used to extract data from unstructured clinical notes has been previously described in detail ^1–4^. The source data were outpatient office notes written by epileptologists at the University of Pennsylvania neurology department. To extract seizure frequencies we fine-tuned pretrained Bio_ClinicalBERT and RoBERTa transformer language models on 700 human-annotated clinical notes. These models classified patients as seizure-free or not since the most recent office visit, and extracted the span of text that contained their seizure frequency. We then used a combination of T5 language model summarization and custom rules-based quantification to convert the extracted text spans into quantitative frequencies. Model performance was comparable to the interrater agreement of 15 human annotators: human F1 = 0.85, model F1 = 0.85. Patients documented as being seizure free were assigned a seizure frequency of zero for that visit. If no seizure frequency was documented for a visit, that visit was excluded from calculating the patient-level estimate of seizure frequency.

For epilepsy diagnosis, focal versus generalized classification, and focal localization, we used DeepSeek-R1, a generative autoregressive language model ^5^. We used few-shot prompting, which supplies a small number of representative input-output pairs as part of the task instructions. Prompts were iteratively refined through cycles of testing and analysis. Model performance was measured on 309 notes annotated independently by 3 expert epileptologists, and the model performance was comparable to human interrater agreement.

Epilepsy type was classified using both six-category (“six-way”) and detailed (“all-way”) schemas. The six-way categories were: focal, generalized, combined generalized and focal, unclassified/unspecified, uncertain if epilepsy, and non-epileptic seizure disorder. Validation performance was: human F1 = 0.82 (SD 0.07) and DeepSeek-R1 F1 = 0.78 (SD 0.02).

The all-way schema provided finer subtype information. Focal epilepsy was further classified as temporal, frontal, parietal/occipital, multifocal, other localized, or unlocalized. Generalized epilepsy was classified as juvenile myoclonic epilepsy, childhood or juvenile absence epilepsy, generalized tonic–clonic seizures alone, Lennox–Gastaut syndrome, or unspecified generalized epilepsy. An additional unspecified category included combined focal and generalized epilepsy, unspecified epilepsy, uncertain if epilepsy, and non-epileptic seizure disorder. Validation performance for all-way typing was: human F1 = 0.72 (SD 0.06) and DeepSeek-R1 F1 = 0.63 (SD 0.005). Full validation results are described elsewhere ^4^.

All analyses were restricted to patients with epilepsy, defined using the six-way classification. Patients were included if their epilepsy type was classified as focal, generalized, combined generalized and focal, or unclassified/unspecified. Patients classified as uncertain if epilepsy or non-epileptic seizure disorder were excluded.

For subtype analyses, patients were classified as having temporal lobe epilepsy or frontal lobe epilepsy based on the all-way classification. Among the remaining patients, generalized epilepsy was assigned if the six-way classification was generalized, excluding combined generalized and focal.

*Supplemental test for state dependence of the spike-seizure correlation*

To assess whether the association between interictal spike rate and seizure frequency depends on clinical state at the time of evaluation, we examined whether spike–seizure correlations were stronger for clinic visits occurring closer in time to EEG recordings.

For each clinic visit, we computed the minimum absolute time difference between the visit date and any EEG recorded for that patient. This visit–EEG interval accounts for patients with multiple EEG recordings (N = 334/3245 patients, 10.3% of the cohort) and aligns each clinic visit with the temporally closest electrophysiologic assessment. Although most patients had only one EEG, 2425/3245 (74.7%) had multiple clinic visits with documented seizure frequency, enabling comparison across visits with differing temporal proximity to EEG.

Visits were stratified by tertiles of the visit–EEG interval distribution, with visits in the lowest tertile classified as short-gap and those in the highest tertile as long-gap. Visits in the middle tertile were excluded to increase separation between groups and reduce misclassification of visits with intermediate temporal proximity. Within each group, patient-level seizure frequency was estimated as the mean reported frequency across visits in the corresponding window, and patient-level interictal spike rate was defined as the mean spike rate per hour across all EEGs for that patient. Analyses were restricted to patients with non-missing seizure frequency and spike rate estimates in both short-gap and long-gap conditions (1147/3245 patients, 35.3%).

We quantified the association between spike rate and seizure frequency separately for short-gap and long-gap visit windows using Spearman correlation. To formally test whether the spike–seizure correlation was stronger for short-gap than long-gap visits, we performed a patient-level bootstrap with resampling of patients with replacement (5,000 iterations). For each bootstrap sample, Spearman correlations were recomputed for short-gap and long-gap conditions, and the difference in correlation coefficients (Δρ = ρ_short gap_ − ρ_long gap_) was calculated. Statistical significance was assessed using a one-sided bootstrap test, defined as the proportion of bootstrap iterations in which Δρ ≤ 0, reflecting the a priori hypothesis that spike–seizure correlations would be stronger for visits temporally closer to EEG.

**SUPPLEMENTAL TABLES AND FIGURES**

| **Model** | **Term** | **Estimate** | **CI_lower** | **CI_upper** | **p** |
| --- | --- | --- | --- | --- | --- |
| Primary (with interaction terms):  *HasSz ~ LogSpikeRate × \|Lag\| + LogSpikeRate × LagDirection + EpilepsySubtype + (1\|Patient)* | | | | | |
|  | Log spike rate | 1.105 | 1.053 | 1.159 | <0.001 |
|  | Absolute lag (years) | 0.935 | 0.889 | 0.979 | 0.002 |
|  | Lag direction (after vs before) | 0.392 | 0.318 | 0.478 | <0.001 |
|  | Frontal vs Temporal epilepsy | 0.686 | 0.432 | 1.094 | 0.11 |
|  | Generalized vs Temporal epilepsy | 0.49 | 0.345 | 0.689 | <0.001 |
|  | Log spike rate x Absolute lag | 0.983 | 0.967 | 0.999 | 0.03 |
|  | Log spike rate x Lag direction | 1.02 | 0.965 | 1.072 | 0.49 |
| Alternate (without interaction terms):  *HasSz ~ LogSpikeRate + \|Lag\| + LagDirection + EpilepsySubtype + (1\|Patient)* | | | | | |
|  | Log spike rate | 1.07 | 1.043 | 1.102 | <0.001 |
|  | Absolute lag (years) | 0.94 | 0.892 | 0.992 | 0.02 |
|  | Lag direction (after vs before) | 0.39 | 0.317 | 0.478 | <0.001 |
|  | Frontal vs Temporal epilepsy | 0.686 | 0.431 | 1.107 | 0.12 |
|  | Generalized vs Temporal epilepsy | 0.492 | 0.346 | 0.697 | <0.001 |

**Supplemental Table 1.** Fixed effects from logistic mixed-effects models predicting seizure occurrence at clinic visits. All models were fit on a table of EEG–visit pairs constructed by cross-joining each patient's outpatient EEGs with their outpatient clinic visits, restricted to patients with a documented epilepsy subtype (temporal lobe, frontal lobe, or generalized; N=12,281 pairs from 1,722 patients). The outcome was whether the patient reported having seizures at the clinic visit (binary). Log spike rate is the natural log of spike rate (spikes/hour) plus an offset of 0.001. Absolute lag is the absolute value of the difference in days between EEG date and visit date, converted to years. Lag direction is coded 1 if the visit occurred on or after the EEG date and 0 if before. Epilepsy subtype effects are referenced to temporal lobe epilepsy. The primary model includes interaction terms between log spike rate and absolute lag, and between log spike rate and lag direction; the alternate model includes the same predictors as main effects only. A likelihood ratio test comparing the two models confirmed that the interaction terms jointly improved fit (χ²(2), p<0.001). All estimates are odds ratios. Confidence intervals and p-values are from cluster bootstrap (5,000 iterations, resampling whole patients).

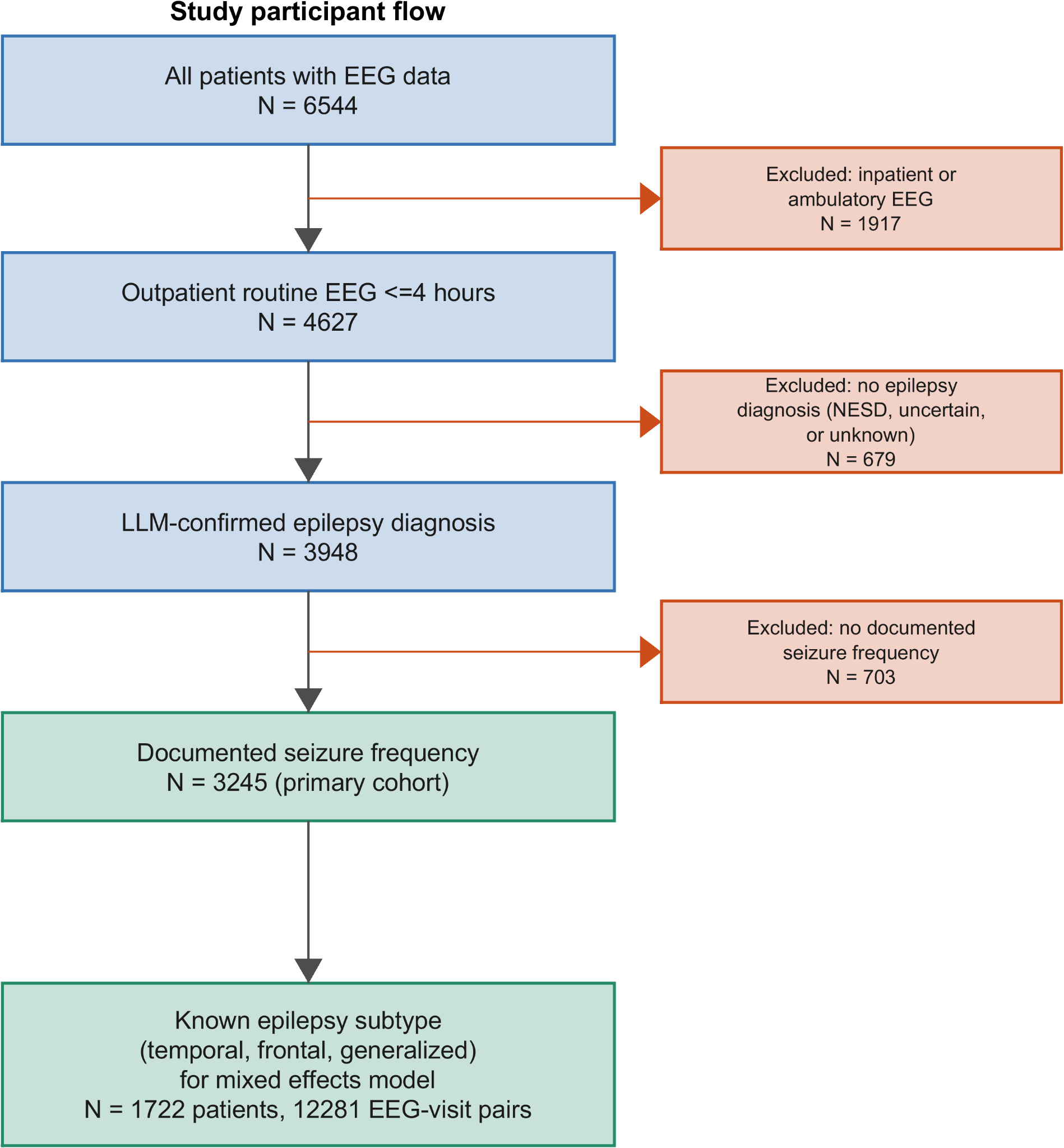

**Fig. S1. Study participant flow diagram.** Starting from all patients with at least one EEG recording in the Penn Epilepsy Center database (N=6,544), patients were excluded sequentially if their EEG was not an outpatient routine recording of less than 4 hours duration (N=1,917 excluded), if they did not carry an LLM-confirmed epilepsy diagnosis (N=679 excluded; includes patients classified as non-epileptic seizure disorder, uncertain if epilepsy, or unknown), or if they had no documented seizure frequency across any outpatient clinic visit (N=703 excluded). The remaining 3,245 patients with epilepsy and at least one documented seizure frequency constitute the primary cohort used for all descriptive and correlation analyses. Of these, 1,722 patients with a known epilepsy subtype (temporal lobe, frontal lobe, or generalized) contributed 12,281 EEG–visit pairs to the logistic mixed-effects model.

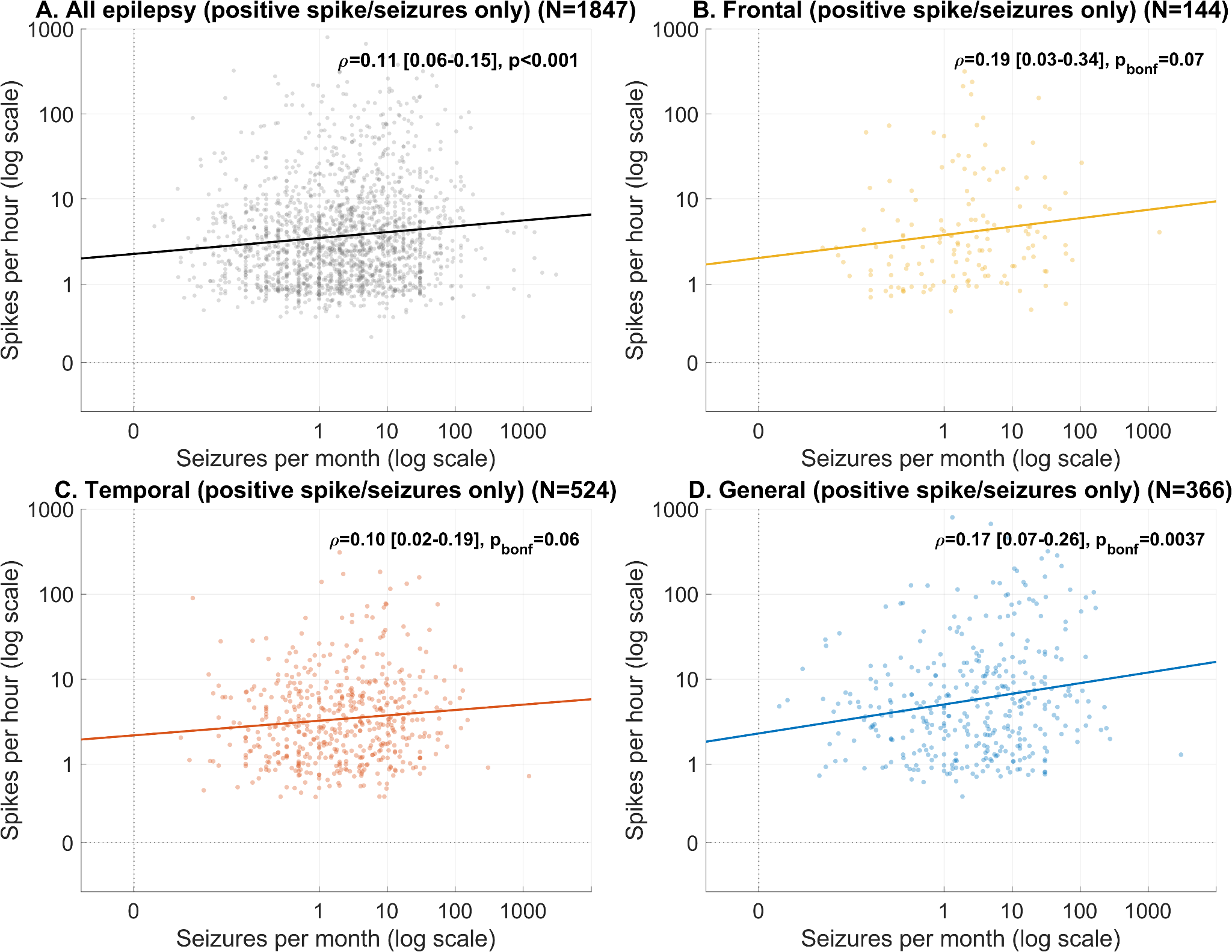

**Fig. S2**. Spike-seizure correlations restricted to patients with positive spike rates and seizure frequencies. This figure shows the results of the same analysis as Figure 3, but restricted to patients with non-zero spike rates and non-zero seizure frequencies.

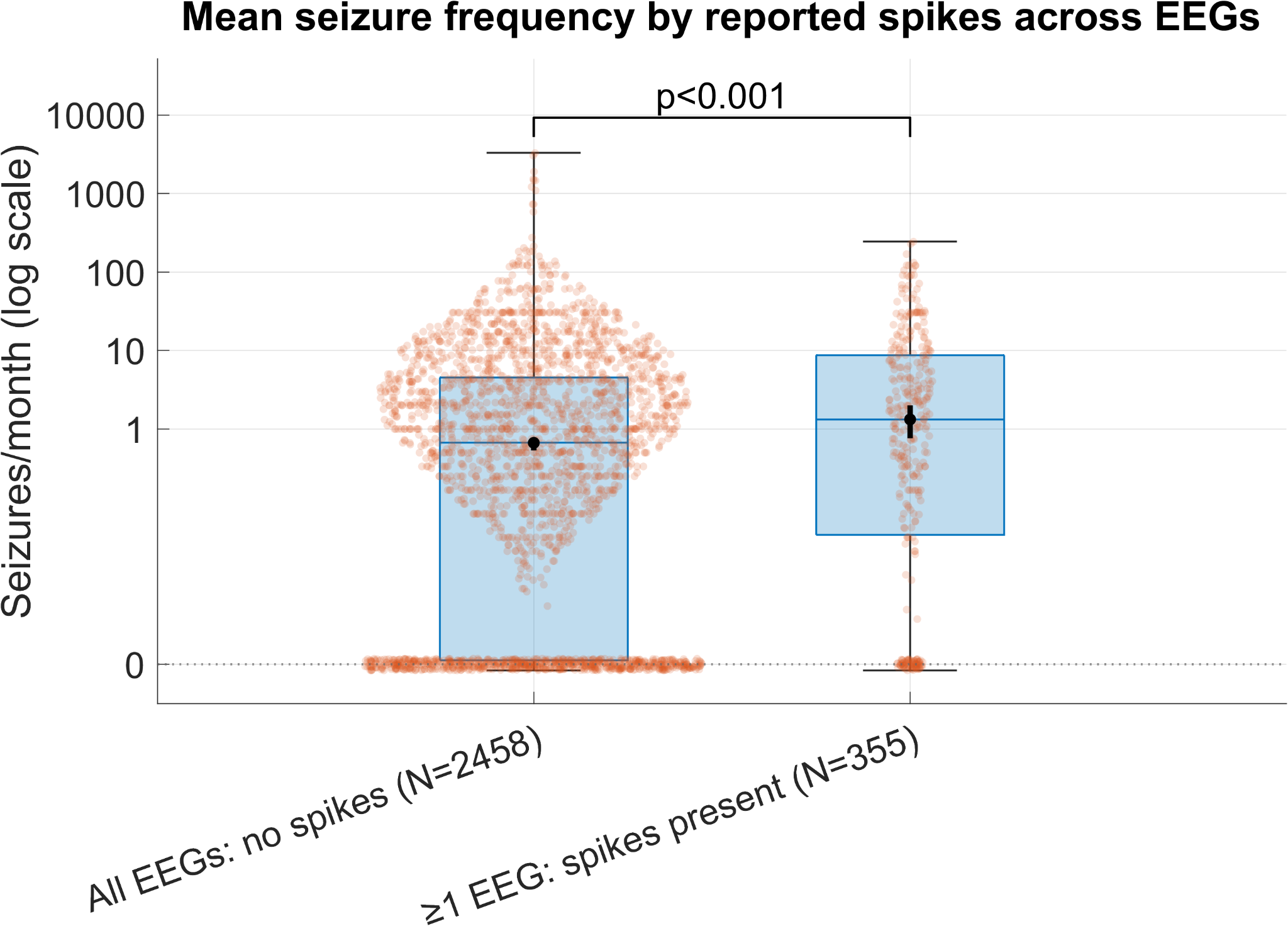

**Fig. S3**. Mean seizure frequency among patients with versus without reported spikes across EEGs. Mean seizure frequency (seizures/month, log₁₀ scale) for epilepsy patients grouped by whether any of their outpatient routine EEGs were reported to contain epileptiform discharges. The dotted line at 0 seizures/month reflects patients with 0 seizure frequency, arbitrarily offset from other points on the log scale. The median [95% CI] seizure frequency was 0.67 [0.54-0.78] seizures/month for patients whose EEGs had no reported spikes and 1.33 [0.76-2.00] seizures/month for patients whose EEGs had reported spikes (p < 0.001, Cliff's δ = 0.12).

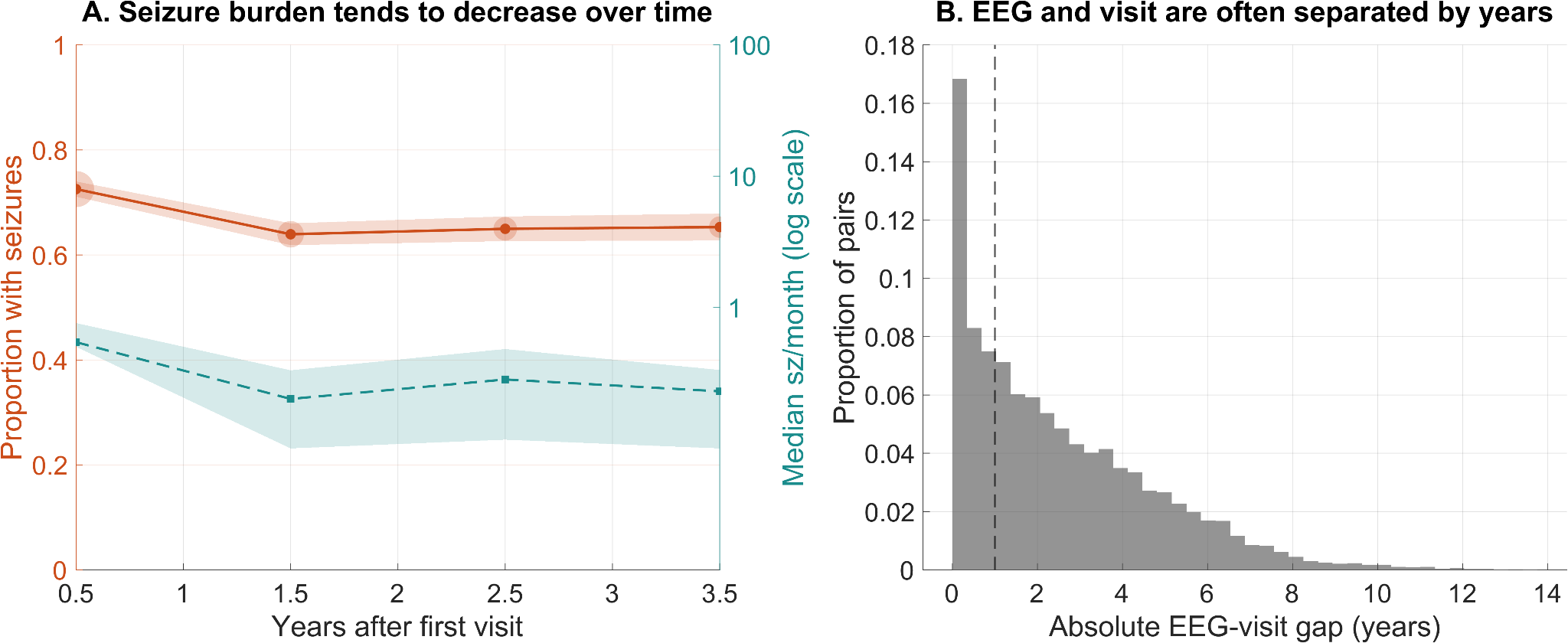

**Fig. S4.** Temporal context for the EEG–visit pair analysis. **A:** Proportion of clinic visits at which seizures were reported (left y-axis, orange) and median seizure frequency in seizures/month (right y-axis, teal), binned by year since first visit. Points represent annual bin means or medians; shaded regions show 95% bootstrap confidence intervals. Marker size is proportional to the number of visits in each bin. The decreasing trend is consistent with a treated epilepsy cohort in which seizure burden improves over time with ongoing care, and motivates the inclusion of visit timing as a covariate in the primary model. **B:** Distribution of absolute EEG–visit lag (in years) across all EEG–visit pairs used in the mixed-effects model (N=12,281 pairs). Each patient contributes one pair per EEG × clinic visit combination. The dashed vertical line marks a 1-year lag. The majority of pairs involve EEGs and visits separated by more than one year, reflecting the real-world clinical pattern in which a single routine EEG is used to inform care across many subsequent visits.

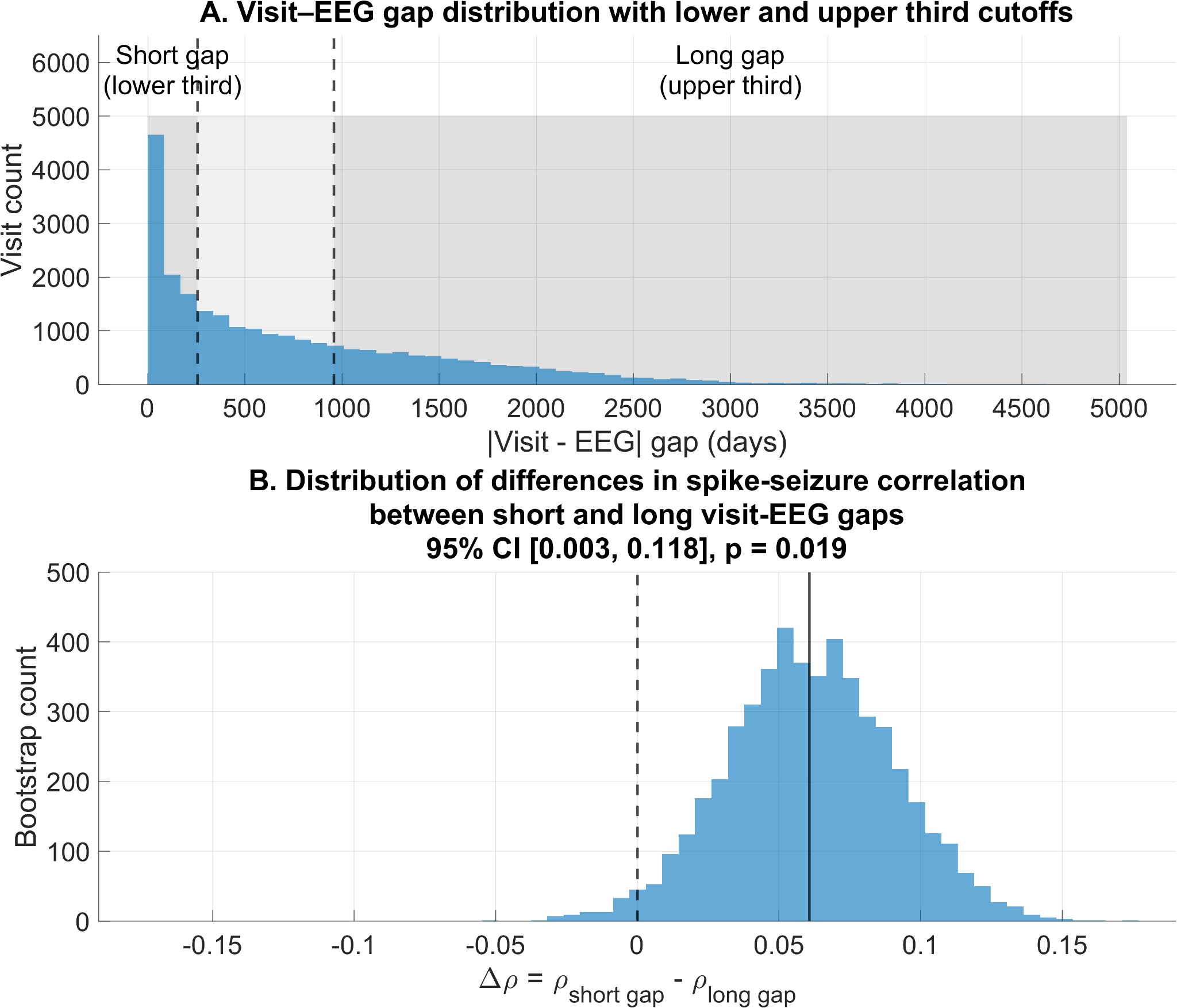

**Fig. S5.** The association between interictal spike rate and seizure frequency is higher for clinic visits close in time to EEGs. A: Distribution of the absolute time difference between clinic visits and EEG recordings, taking the minimum in the case of multiple EEGs per patient. Visits were stratified into short gap and long gap groups based on tertiles of the visit–EEG gap distribution (lower third (256 days) = short gap; upper third (959 days) = long gap). Shaded regions indicate the lower, middle, and upper thirds of the gap distribution, with dashed vertical lines marking the tertile cutoffs (256 days and 959 days). B: Bootstrap distribution (5000 iterations) of the difference in Spearman correlation coefficients between interictal spike rate and seizure frequency for near versus far visit windows (Δρ = ρ_short gap_ − ρ_long gap_). The spike–seizure correlation was stronger when clinic visits occurred closer in time to EEG acquisition (N = 1147 patients with both short-gap and long-gap visits; ρ_short gap_ = 0.15; ρ_long gap_ = 0.09; observed [95% CI] Δρ = 0.061 [0.00–0.12], one-sided p = 0.019).
